# Impact of cell type specific variations and age in aortic distensibility

**DOI:** 10.1101/2025.11.25.25340959

**Authors:** Mehak Chopra, Niamh Hynes, Cathal Seoighe

## Abstract

Aortic distensibility refers to the ability of arteries to expand in response to pulse pressure generated by the cardiac cycle, and this often decreases with age. Genome-wide association studies have identified genetic variants associated with distensibility; however, the mechanisms leading to changes in distensibility remain unclear. In this study we examined aortic distensibility through the lens of genomics, considering both cellular composition and cell type specific gene expression, inferred from bulk gene expression data, to investigate how these factors contribute to the observed changes in distensibility associated with age and genotype. We found age-related decreases in the proportions of Pericytes and Fibroblast I cells, while the proportion of vascular smooth muscle cells type II (VSMC II) increased. Notably, most of the gene expression changes associated with age were identified in VSMC I, VSMC II and Fibroblast I cells. Furthermore, we observed that the cell type-specific expression of most genes associated with distensibility correlated with age, specifically VSMC I, VSMC II, Fibroblast I, and Pericyte cells. We also tested for genetic associations with the extent of increased distensibility with age in the UK Biobank and found two independent loci, both of which showed a marginally significant association with the increased distensibility with age. None of the identified GWAS SNPs were significantly associated with the inferred cellular proportions. Interestingly, we found two independent SNPs that had a genome-wide significant association with distensibility were also associated with cell type specific ex-pression of nearby genes (*SRR* in VSMC I, VSMC II and Fibroblast I, as well as *CDH13* in VSMC I) that have been implicated in aortic distensibility. Overall, our results identify cell type specific changes in gene expression that may help to explain genetic and age-related variation in this important physiological phenotype.

## Introduction

The aorta, the largest artery in the body, carries oxygen-rich blood from the left ventricle through the aortic valve to the circulatory system. The elasticity of the aorta allows it to expand, enabling it to accommodate half of the blood volume ejected on each beat, effectively acting as a blood reservoir [1, 2]. Aortic elasticity is described by a parameter known as distensibility, which is a measure of the extent to which the aorta expands in response to the pulse pressure induced by cardiac contraction and relaxation [3]. A decrease in distensibility may impair the ability of the aorta to moderate the high pulsatile pressure of the left ventricle, exposing the cardiovascular system to increased stress. This heightened stress can lead to cardiovascular abnormalities [2], adversely affecting the normal functioning of the left ventricle, heart, and blood flow [4].

The aortic wall is composed of several extracellular matrix components such as elastin fibres and collagen and multiple cell types, including vascular smooth muscle cells (VSMCs) and fibroblasts. With advancing age, the composition of the tissue may change, contributing to the phenotypic characteristics associated with cardiovascular ageing. For example, the VSMCs stiffen while elastin fibres tend to rupture, leading to diminishing distensibility over time [5]. Some other factors like hypertension, hyperlipidemia and genetics as well as external factors like smoking are also associated with stiffening of the aorta, which could lead to acute aortic syndromes (AAS) and aortic aneurysms [6].

Distensibility can be accurately obtained using Cardiovascular Magnetic Resonance Imaging (CMRI), which can predict adverse cardiovascular events, unravelling the relationships between imaging phenotypes and aortic disorders. Large-scale imaging investigations may help yield information in the search for disease-risk factors and early-stage image-based biomarkers [7]. Studies such as the Multi-Ethnic Study of Atherosclerosis (MESA), which enrolled 6,814 healthy participants from six American communities between the ages of 45 and 84, can help to determine the causes of cardiovascular disorders [8]. The Dallas Heart Study, which had 3,000 participants [9]; the Jackson Heart Study, with 2,000 participants [10]; and the Framingham Heart Study, which began in 1948 and has over 15,000 participants over three generations, are among the studies that demonstrate the potential of CMRIs. However, due to the small number of participants enrolled compared to large-scale studies like the UK Biobank (UKB), such studies may lack the power to explore the genetic variations associated with a wide range of phenotypes. The UKB is a large cohort of approximately 500,000 middle-aged and older adults recruited between 2006 and 2010 [11]. It includes imaging, genotypic and rich phenotypic and clinical data. Many genome wide association (GWA) studies have utilized data from UKB to estimate the association of single nucleotide polymorphisms (SNPs) with phenotypes [12]. Previous GWA studies of aortic distensibility using data from UKB have identified several associated genetic variants and genes, supporting a genetic contribution to variation in aortic elasticity [13]. While distensibility varies among individuals, with ageing being the primary factor contributing to reduced aortic distensibility, genetic effects could either accelerate or slow down this process [14]. Despite the established genetic associations, the mechanisms driving changes in distensibility remain unclear.

In this study, we investigated the mechanisms underlying variation in distensibility from a genomics perspective. Our hypothesis was that both variation in the cellular composition of the aorta and variation in gene expression within specific cell types may contribute to the observed variation in distensibility with age and by genotype. We first applied gene expression deconvolution [15] to bulk aortic gene expression data from the GTEx consortium [16] and examined the changes in cell type proportions with age as well as cell-type specific changes in gene expression with age. We then performed a GWA of distensibility in 47,674 samples from the UKB for which cardiac magnetic resonance imaging (MRI) data were available and investigated the relationship between genetic variants associated with distensibility and inferred cell type proportions, as well as performing cell type-specific gene expression (CT-GE) analysis to determine whether gene expression changes in specific cell types may potentially mediate the identified genetic effects on aortic distensibility. Although a GWA study of distensibility has been carried out previously that uses the majority of these samples, additional analyses reported here shed light on the mechanisms of genetic variation in distensibility and its relationship to the decrease in distensibility seen with age. Our results suggest that the cellular composition of the aorta changes with age and that changes in the expression of key genes in vascular smooth muscle cells and fibroblasts may explain some of the genetic associations of aortic distensibility.

## Data and methods

### Distensibility from CMRIs

Genotypic and phenotypic data were accessed from UKB under application number 74519. These included cardiac MRI (CMRI) data, which was generated by UKB using a wide bore 1.5 Tesla scanner (MAGNETOM Aera, Syngo Platform VD13A, Seimens Healthcare, Erlangen, Germany) [17, 18]. Steady-state free precession cine aortic CMRIs taken at the level of the right pulmonary trunk were obtained in DICOM (Digital Imaging and Communications in Medicine) format from the UKB. A set of 100 DICOM image sequence frames covering the cardiac cycle for each participant had a typical size of 240 *×* 196 pixels with a spatial resolution of 1.6 *×* 1.6 mm^2^. We employed an existing fully automated method of image segmentation on 62,497 CMRIs, which uses a convolutional neural network developed by Bai, Wenjia, et al. (2018) [19] employing CUDA (v11.8.0) and cuDNN (v8.6.0) with Tensorflow (v2.12.1). 1,651 CMRIs were excluded after applying quality checks described by Bai, Wenjia, et al. (2020) [7]. The largest (*A*_max_) and the smallest areas (*A*_min_) of both aortic regions were calculated. Using the mean systolic (SBP) and diastolic blood pressure (DBP) data for the same participants we then calculated the ascending and descending distensibility for 56,765 CMRIs with the following equation:

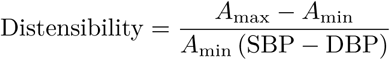

### Cellular deconvolution and cell type specific gene expression

Aortic gene expression and phenotypic data from the GTEx consortium (V8), including age and sex, were obtained from the GTEx Portal [16]. Cellular proportions in bulk gene expression data from 432 GTEx aortic samples were estimated using CIBERSORTx [20]. Ensembl gene IDs were converted to gene symbols using biomaRt [21] and Ensembldb [22] R packages after removing duplicate genes. Using Seurat and Seurat-disk [23] R packages, a cell-type gene reference matrix was derived from a single-nucleus RNA sequencing study of ascending aortic tissue samples with GEO accession ID GSE165824, which included 54,092 nuclei and 12 identified primary cell clusters [24]. Gene expression deconvolution was performed using s-batch. In addition to cellular composition, cell type-specific gene expression was estimated using CIBERSORTx high-resolution mode on the bulk gene expression data with the gene reference matrix described above.

### Functional enrichment analysis

The correlation between cell type-specific gene expression and age was assessed by calculating the Spearman correlation coefficient in R. Utilising the estimated correlation values and standard error derived from the R package statpsych [25], we conducted gene set analysis using a non-parametric approach using the Retest function of mremaR [26]. A second gene set analysis was performed using fgsea [27] by ranking the genes by correlation values. Overrepresentation analysis was also performed using significant positively and negatively correlated genes with age as input, respectively, and all correlated genes per cell type as the universe in fgsea fora. For all three gene set analysis methods, we utilized hallmark gene sets (H), canonical pathways (CP), gene ontology gene sets (GO), and human phenotype ontology (HPO) gene sets from the GSEA human molecular signatures database (MSigDB) [28–30].

### Genome wide association analysis and Heritability

Following previous studies on aortic dimensions, such as aortic diameter and aortic distensibility by Pirruccello, James P et al. [24], Francis, Catherine M et al. [13] respectively, the genotypic data for 55,247 individuals was filtered by minor allele frequency (threshold 0.001), missing call rates (0.02) and Hardy-Weinberg equilibrium p value (with mid p-adjustment and a threshold 5*×*10*^−^*^7^). Correlated SNPs were pruned using a window size of 50, step size of 5, and a correlation of 0.2. Participants exhibiting aortic distensibility exceeding four standard deviations above the mean were included in the analysis. GWAS on ascending and descending distensibility was performed using a Bayesian linear mixed model tool called BOLT-LMM (v2.4.1) [31, 32] with a genetic map file accounting for recombination scores and previously estimated LD scores (https://alkesgroup.broadinstitute.org/Eagle/downloads/tables/) for 1000 genomes available with BOLT. We ran BOLT-LMM on 47,674 white British and Irish participants with --predBetasFile flag to estimate the SNP beta coefficient for polygenic prediction. To minimise background genetic noise, covariate adjustment was employed with genotypic batch and MRI centre, which were kept as qualitative covariates. Data from participants with extreme Body Mass Index (BMI) values (i.e., *<* 15 or *>* 40) were excluded. To minimize the influence of extreme blood pressure values, we excluded individuals with highly elevated hypertension measures, DBP *>* 100 mmHg or SBP *>* 160 mmHg (i.e., stage 2 hypertension) [33]. Age, sex, anthropometric traits like height, weight, BMI, systolic and diastolic blood pressure, and the top 10 principal components were included as quantitative covariates. Estimation of the components of heritability was carried out using BOLT-REML (v2.4.1) on each chromosome.

### Gene-based analysis

The lead SNPs and independent significant SNPs, associated with aortic distensibility were identified using FUMA (v1.5.2) SNP2GENE [34]. The maximum p value of lead SNPs was kept at *<* 5e-8, with a maximum p value cutoff of *<* 0.05. The independent significant SNPs were defined using the r2 threshold of 0.6, whereas the r2 threshold of 0.1 was used for lead SNPs. The maximum distance between LD blocks to merge into a locus was *<* 250 kb. The maximum distance was kept at 10 kb for gene positional mapping. eQTL mapping was performed using only significant SNP-gene pairs based on aortic samples in GTEx v8 with FUMA SNP2GENE (FDR 0.05 and p value *<* 1e-3). 2D Chromatin interaction mapping was performed based on HiC data from aorta (GSE87112) within a promotor region window of 250 bp upstream and 500 bp downstream from the transcriptional start site and FDR threshold *<* 1e-6 for significant loops. Enhancer and promoter regions were annotated using the aorta epigenome (E065) from the Roadmap Epigenome Project [35]. We used MAGMA [36] to perform a gene-property analysis testing with tissue-specific gene expression levels on 54 tissue types in GTEx V8 with GWAS gene-level association statistics.

### SNP-age interaction and GTEx genotypic data analysis

We tested for a statistical interaction between the genotype of distensibility-associated SNPs and age in UKB using linear statistical modelling, treating distensibility as the response and SNP-age interaction as a predictor, including sex as a covariate and correcting for multiple testing using the Benjamini-Hochberg method.

We selected independent significant SNPs that showed an association with distensibility in the UKB GWAS and conducted a linear regression analysis using the R package RegParallel [37] to test whether any of these SNP genotypes were significantly associated with the estimated cellular proportions, controlling for age and sex, following the Benjamini-Hochberg (BH) method. For the genes implicated in the GWAS we tested for an association between their expression level in each of the aortic cell types and the genotype of the independent SNPs while adjusting for age and sex. For this analysis we used the gene expression levels inferred using CIBERSORTx and again performed correction for multiple testing using the BH method.

## Results

### The decrease of aortic distensibility with age

We utilized the image segmentation model and pulse pressure outlined in the Methods section to assess the ascending and descending distensibility for a total of 56,765 UKB participants. As expected, given previous studies [13, 38], aortic distensibility tends to decrease with age. The proportion of variation in the normalized ascending and descending distensibility in this cohort explained by age after adjusting for sex was 0.256 and 0.259, respectively (Figure 1).

**Figure 1:**
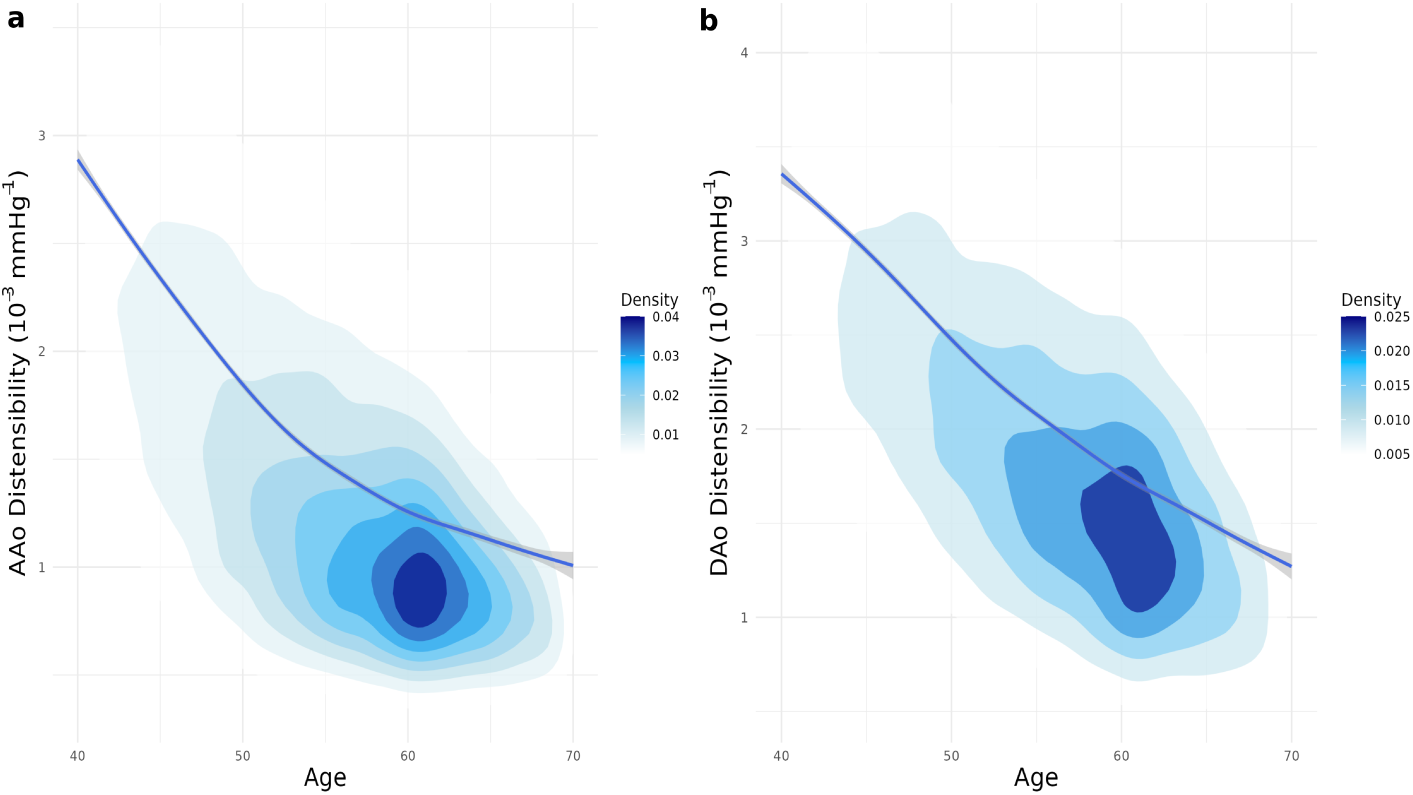
a,b) Inverse association between age and aortic distensibility. Den-sity plots showing aortic distensibility (in units of 10*^−^*^3^ mmHg*^−^*^1^) as a function of age (years). A negative association (p value *<* 2.2 *×* 10*^−^*^16^) is observed, indi-cating reduced ascending and descending distensibility with advancing age.

### Change in cellular composition of aorta with age

Using cell type deconvolution, we estimated the proportions of 12 different cell types in 432 GTEx aortic tissues samples [Figure 2-a]. As distensibility declines with age, we sought to investigate whether there are corresponding changes in the cellular composition of the aorta with age. Pericytes and fibroblast I exhibited a significant decline with age (BH-adjusted p value of 2.1e-11 and 2.9e-04, respectively; Figure 2b-c) while vascular smooth muscle cells type II showed a significant increase with age after controlling for sex (BH-adjusted p value of 1.1e-05; Figure 2-d). These associations suggest that age-related changes in the cellular composition of the aorta may make a contribution to changes in aortic distensibility with age.

**Figure 2:**
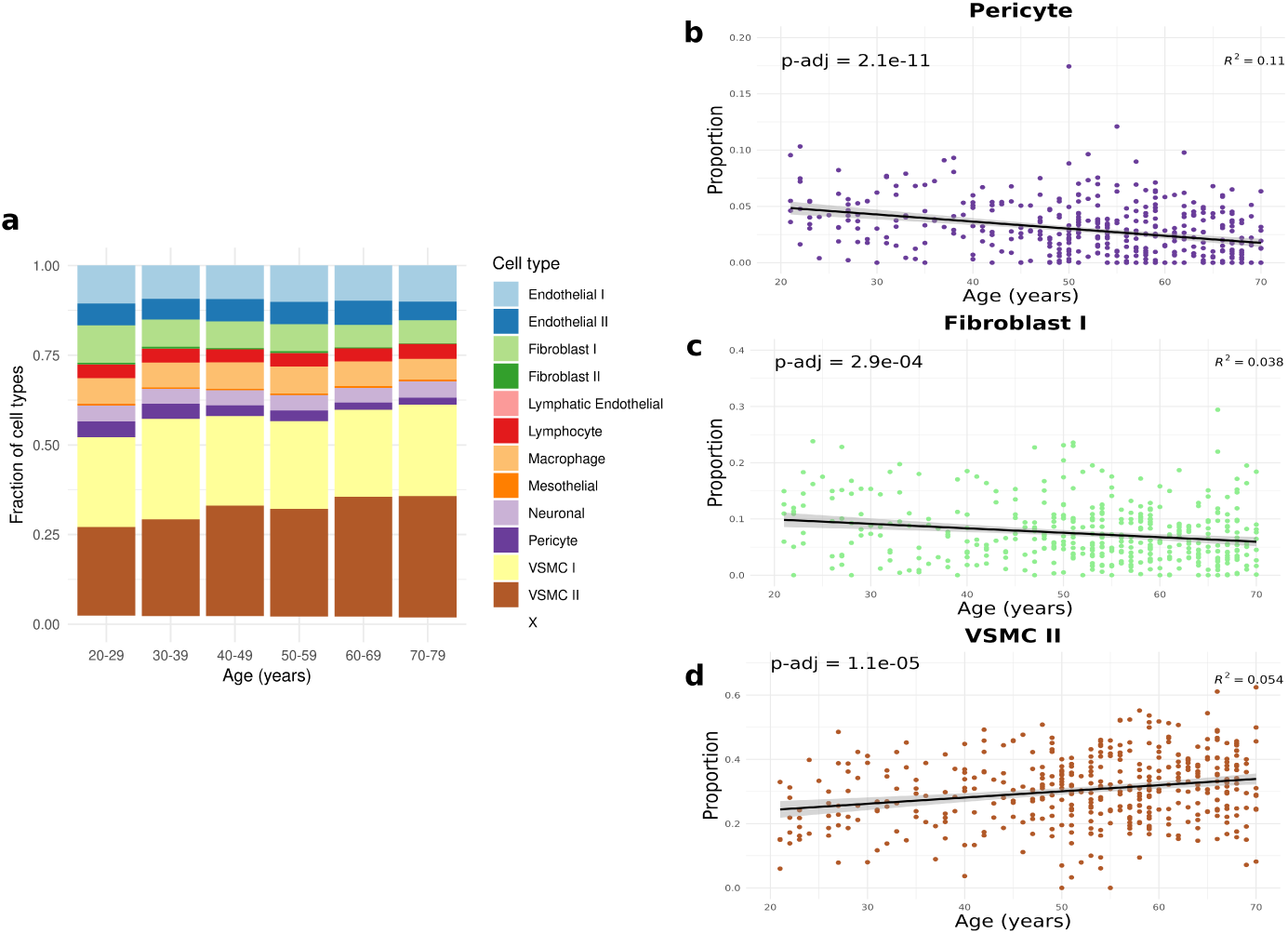
a) Cellular composition of 432 GTEx aorta samples, estimated using gene expression deconvolution. b-d) Scatterplots of cellular proportions as a function of age for the three cell types Pericyte, Fibroblast I, VSMC II that showed changes in proportion with age (BH corrected p value (p-adj) after ad-justing for sex). The linear model fit and 95% confidence envelope with *R*^2^ values are shown for each linear model fit.

### Evaluating age-related gene expression in distinct cell types

Using CIBERSORTx HiRes, we imputed sample-level gene expression for each cell type from the GTEx aortic bulk RNA-seq data. Among the 12 cell types analysed, the majority of genes associated with age were identified in VSMC type I (VSMC I) and VSMC type II (VSMC II), followed by Fibroblast type I (Figure 3). This suggests that many age-related changes in gene expression occur in smooth muscle cells and other cell types, potentially impacting the physical properties of the cells or the extracellular matrix of the aorta.

**Figure 3:**
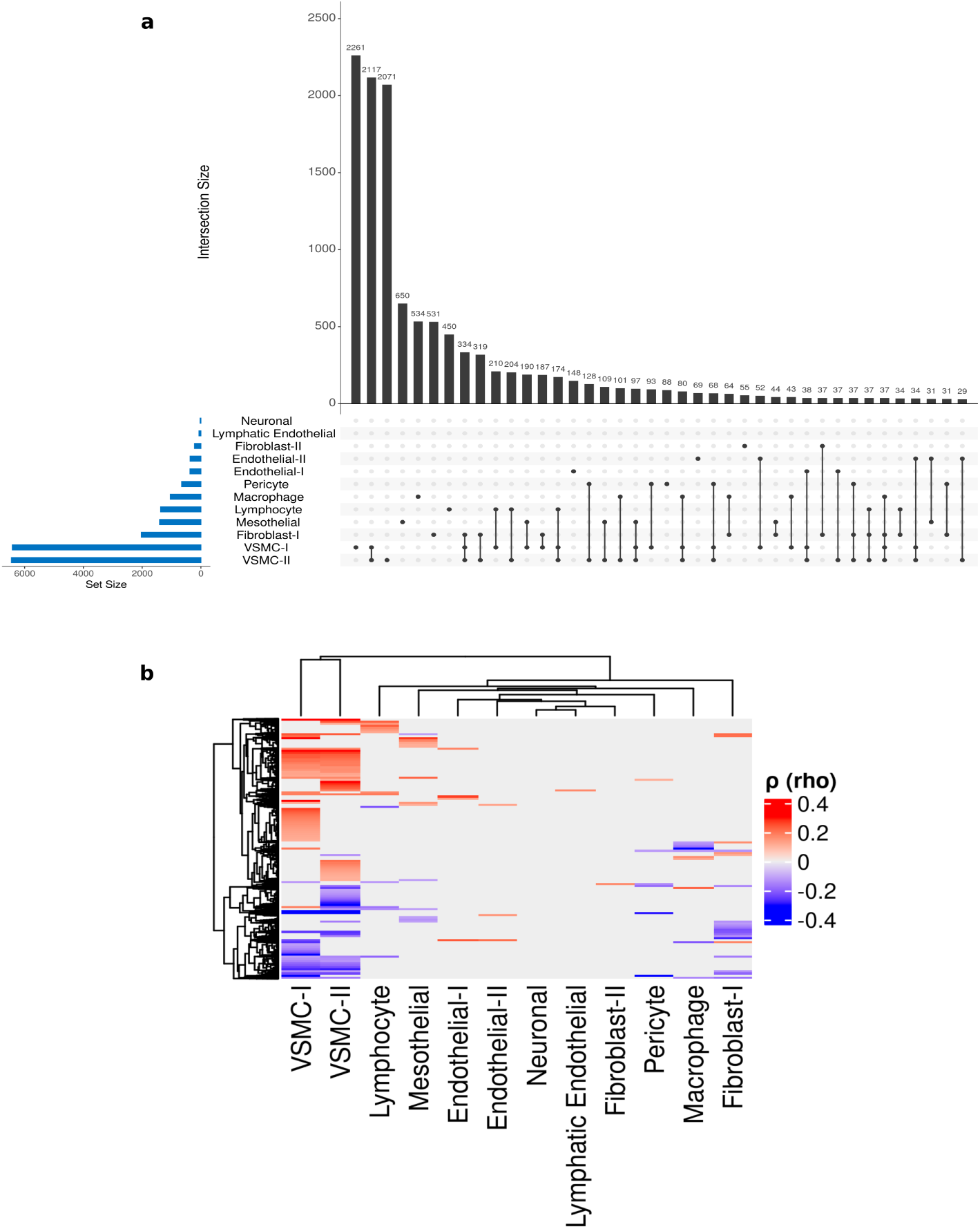
a) UpSet plot showing the intersections between the genes corre-lated with age in specific cell types. The bars on the left represent the number of genes significantly correlated with age in individual cell types. The matrix of connected dots indicates the cell types corresponding to each intersection, and the vertical bars above represent the size of each intersection. b) Heatmap illustrating genes (on the rows) for which expression in the cell types in the columns was significantly positively (red) and negatively (blue) correlated with age.

### Gene set analysis of age-associated genes

We applied three separate gene set analysis methods to explore the genes for which there was an association between cell-type specific expression and age (see Data and Methods). Interestingly, all three methods revealed that genes with expression that was negatively correlated with age were enriched in the HPO gene sets *Abnormality Of Connective Tissue* and *Increased Inflammatory Response* in VSMC-I (Supplementary Figure 1). In the GOBP gene sets, we observed an enrichment in genes associated with the *Regulation of Membrane Potential* in VSMC-I, *Vasculature Development* in VSMC-II, and *Reactome Vesicle-Mediated Transport* gene set in VSMC-I using Reactome pathways (Figure 4). These results are consistent with age-associated changes, concentrated in specific gene sets in VSMC-I and VSMC-II, critical cell types for aortic distensibility, potentially having an influence on the changing properties of the aorta as individuals age.

**Figure 4:**
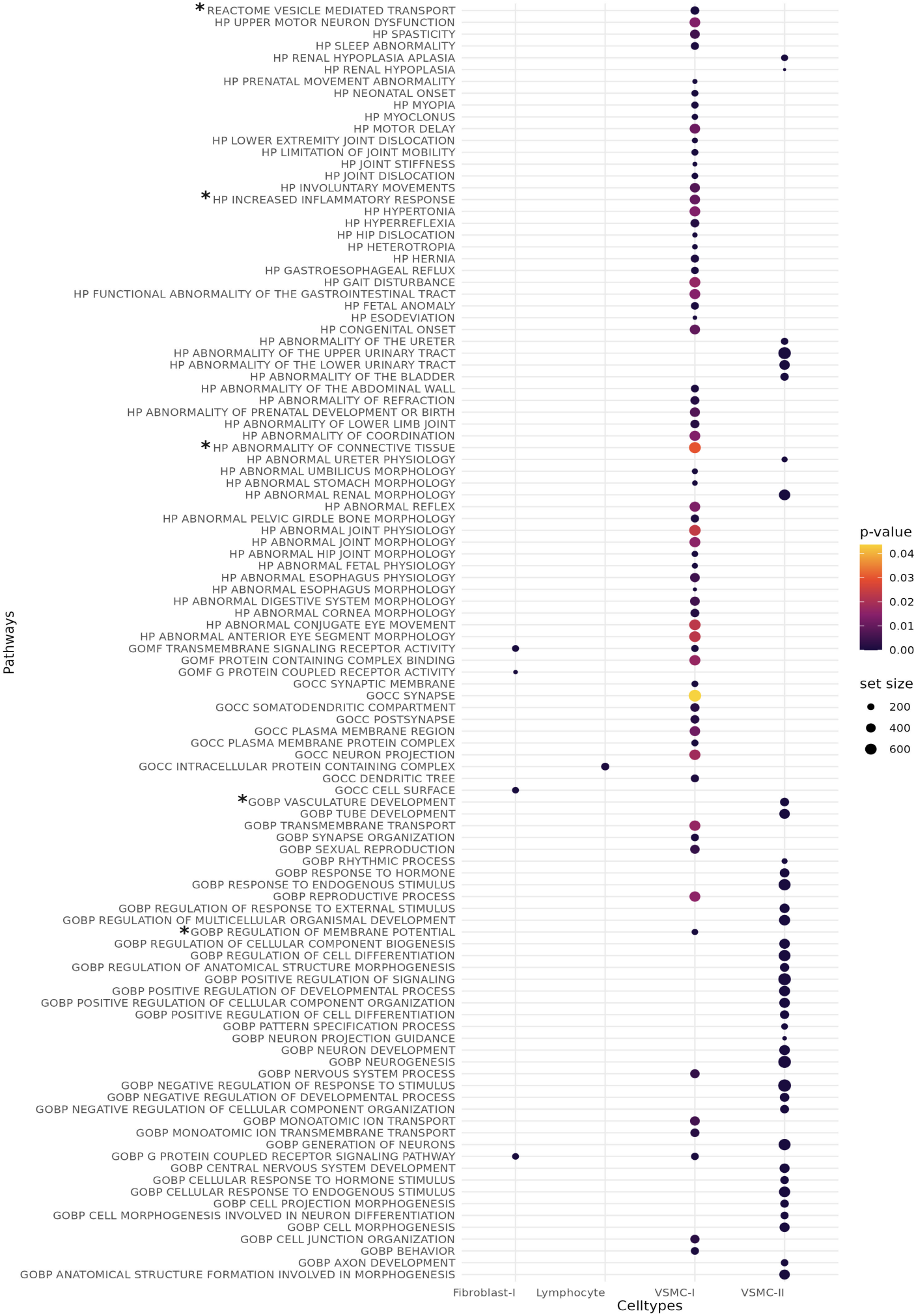
Common gene sets that were significantly enriched for a negative correlation with age were identified in Fibroblast-I, Lymphocyte, VSMC-I, and VSMC-II using three gene set analysis methods (fgsea, fora, and mremaR). The p value, set-size values are from mremaR and asterisk(*) indicates the enriched gene sets that may be associated with the ageing aorta.

### Genome wide association analysis of ascending distensibility

GWA analysis was performed on ascending and descending distensibility using BOLT-LMM and 6,741,306 SNPs that passed QC, accounting for population stratification and cryptic relatedness. For ascending distensibility 1,757 SNPs in 33 independent loci passed the genome-wide significance threshold of 5e-8 (Table 1; for descending distensibility, see Supplementary Figure 2 and Supplementary Table 2). Unsurprisingly, the results we obtained were consistent with those that have been reported in earlier GWA studies that were carried out on the majority of the samples we analysed (32,639 [39]; 29,895 [13]; 29,684; [40]). Using FUMA, the 33 independent loci could be mapped to 27 genes (Figure 5-a). Applying MAGMA, a tool that performs gene set analysis on GWAS results, showed that genes highly expressed in aorta tissue tended to have stronger GWAS signals (p value = 0.0024). The highest SNP heritability estimate was found for chromosome 5, followed by chromosome 16. The overall SNP-heritability of the distensibility of the ascending aorta was estimated to be 0.24, suggesting that genetic factors account for a moderate proportion of the variation in ascending aortic distensibility.

**Table 1:** Significant independent SNPs and mapped genes associated with as-cending distensibility using FUMA.

| Genomic Locus | Independent SNP rsID | Chr | BP | p value | Mapped genes |
| --- | --- | --- | --- | --- | --- |
| 1 | 3:41896441_TAA<br>AAAAAAAAAAAAA_T | 3 | 41896441 | 1.70E-10 | *EIF1B*,<br>*ENTPD3*,<br>*ULK4*,<br>*ZNF621*,<br>*TRAK1* |
| 2 | rs4371706 | 5 | 51185244 | 2.50E-11 |  |
| 3 | rs7447610 | 5 | 95605114 | 6.30E-13 | *ELL2*,<br>*ARSK*,<br>*RHOBTB3*,<br>*GLRX*,<br>*PCSK1*,<br>*RFESD*,<br>*GPR150*,<br>*TTC37*,<br>*C5orf27* |
| 3 | rs4336380 | 5 | 95614860 | 4.00E-11 | *ELL2*,<br>*ARSK*,<br>*RHOBTB3*,<br>*GLRX*,<br>*PCSK1*,<br>*RFESD*,<br>*GPR150*,<br>*TTC37*,<br>*C5orf27* |
| 4 | rs337101 | 5 | 122550646 | 8.00E-12 | *PRDM6* |
| 5 | rs4717861 | 7 | 73416183 | 4.60E-17 |  |
| 5 | rs10224499 | 7 | 73423846 | 2.30E-09 |  |
| 5 | rs11768878 | 7 | 73431169 | 1.00E-31 | *ELN* |
| 5 | rs73144900 | 7 | 73440265 | 8.60E-12 |  |
| 5 | rs1859761 | 7 | 73454629 | 8.50E-10 | *ELN* |
| 5 | rs810549 | 7 | 73493533 | 1.50E-08 | *ELN*,<br>*LIMK1* |
| 6 | rs7831476 | 8 | 75694909 | 4.10E-08 | *PL15*,<br>*RP11-758M4.1* |

| Genomic Locus | Independent SNP rsID | Chr | BP | p value | Mapped genes |
| --- | --- | --- | --- | --- | --- |
| 6 | rs71527276 | 8 | 75757172 | 1.00E-09 | *PL15,*<br>*RP11-758M4.1* |
| 6 | rs141775964 | 8 | 75780655 | 7.40E-11 | *PL15,*<br>*RP11-758M4.1* |
| 6 | rs2732010 | 8 | 75784980 | 1.90E-11 | *PL15,*<br>*RP11-758M4.1* |
| 7 | rs11781907 | 8 | 122638989 | 5.40E-11 | *HAS2* |
| 7 | 8:122674851_TTA_T | 8 | 122674851 | 5.70E-10 | *HAS2* |
| 7 | rs28545768 | 8 | 122675516 | 1.20E-08 | *HAS2* |
| 7 | rs10102879 | 8 | 122681402 | 1.00E-11 |  |
| 8 | rs10086151 | 8 | 124541280 | 6.30E-10 | *FBXO32* |
| 8 | rs10101794 | 8 | 124541764 | 1.10E-08 | *FBXO32* |
| 8 | rs6470156 | 8 | 124579985 | 1.90E-11 | *FBXO32* |
| 8 | rs11776999 | 8 | 124594923 | 2.20E-08 | *FBXO32* |
| 8 | rs34557926 | 8 | 124607159 | 1.50E-12 | *FBXO32* |
| 9 | rs9336085 | 10 | 30083657 | 1.10E-10 | *SVIL* |
| 9 | rs2150562 | 10 | 30162423 | 4.30E-09 | *SVIL* |
| 9 | rs914279 | 10 | 30170487 | 3.50E-12 | *SVIL* |
| 10 | rs11187803 | 10 | 95944250 | 3.00E-08 | *PLCE1* |
| 11 | rs72792127 | 16 | 83022702 | 3.10E-08 | *CDH13* |
| 11 | rs12445776 | 16 | 83036902 | 4.70E-09 | *CDH13* |
| 12 | rs6503302 | 17 | 2052704 | 9.00E-10 | *SMG6,*<br>*SRR* |
| 12 | rs2126202 | 17 | 2095712 | 2.50E-10 | *SMG6,*<br>*SRR* |
| 12 | rs216199 | 17 | 2200871 | 1.80E-11 | *SMG6,*<br>*SRR,*<br>*TSR1* |

**Figure 5:**
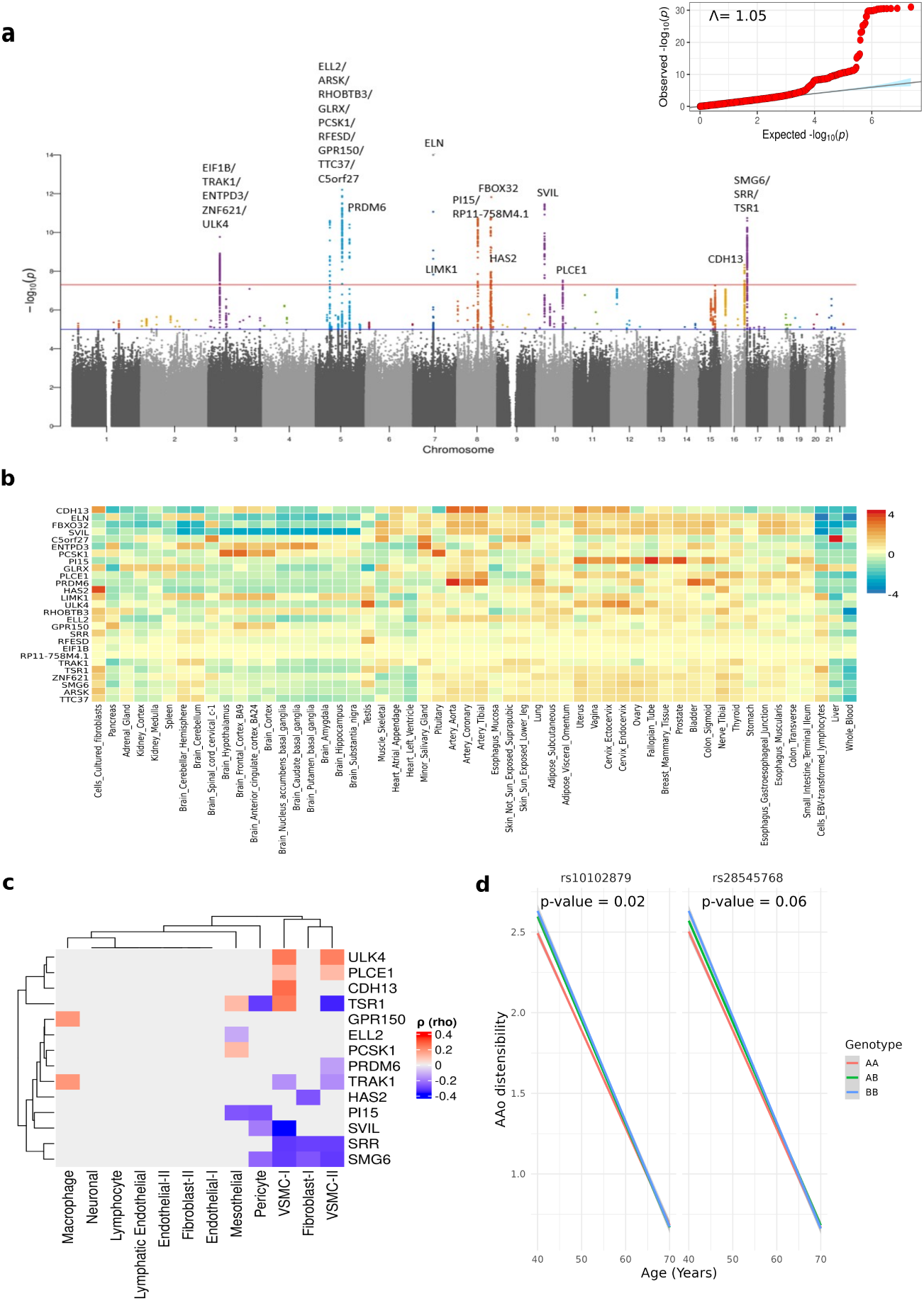
a) Manhattan plot from the GWAS analysis of aortic ascending dis-tensibility. The x axis represents chromosomes 1 to 22 and the y axis represents the negative log (to the base 10) of the two-sided p values for association of vari-ants with ascending aortic distensibility. b) Gene expression heatmap showing normalised expression per GTEx tissue (zero mean across samples) of the 27 genes mapped to significant GWAS loci across 54 GTEX tissues. c) Heatmap il-lustrating the correlation of cell-type specific gene expression (of the 14 mapped genes out of 27 to significant independent SNPs linked to ascending distensibil-ity) with age, highlighting positive correlations in red and negative correlations in blue. d) Genotype-age interaction effect for ascending aortic distensibility for SNPs rs10102879 and rs28545768. Genotypes are categorised as AA, AB, and BB. AA represents the absence of the effect allele, AB corresponds to het-erozygotes, and BB indicates homozygosity for the effect allele (i.e., carrying two copies of the effect allele).

### SNP-age interaction analysis

We investigated whether there was evidence that any of the loci significantly associated with aortic distensibility also affected the extent of increased distensibility with age. To do this, we included the interaction term between SNP and age in the linear model for distensibility. Two SNPs (rs10102879 and rs28545768, both of which are upstream of gene *HAS2* (Hyaluronan synthase 2) showed a marginally significant BH p value of 0.02 and 0.06, respectively (Figure 5-d). This indicates a potential age-dependent genetic effect for variants at this locus.

### Cellular composition as a potential mechanism of genetic association with ascending distensibility

To investigate whether genetic variation in cell-type composition of the aorta may explain some of the genetic variants associated with aortic distensibility, we tested all loci identified in the GWA for an association with the inferred cellular proportions in the GTEx aorta samples. GWA loci rs141775964, rs7831476 and rs10101794 showed nominally significant associations with Fibroblast II, VSMC I and VSMC II proportions, respectively; however, none of these associations were significant following correction for multiple testing using the Benjamini-Hochberg method (Supplementary Table 3, Supplementary Figure 5).

### Cell-type specific eQTL analysis

We tested whether genetic variants associated with distensibility in the UKB were also associated with inferred cell type–specific gene expression in the GTEx aortic samples. Using CIBERSORTx HiReS, we imputed sample-level gene ex-pression for distinct cell types from GTEx aortic bulk gene expression data. Using linear regression, we found that rs6503302 and rs2126202 were associated with variation in *SRR* (Serine Racemase) gene expression in VSMC I, VSMC II, and Fibroblast I (Figure 6a,6c-d). Additionally, rs72792127 and rs12445776 were found to be associated with gene expression of *CDH13* (Cadherin 13) in VSMC-I (Figure 6b). This suggests that altered expression of *SRR* and *CDH13* within these cell types may contribute to genetic variation in distensibility.

**Figure 6:**
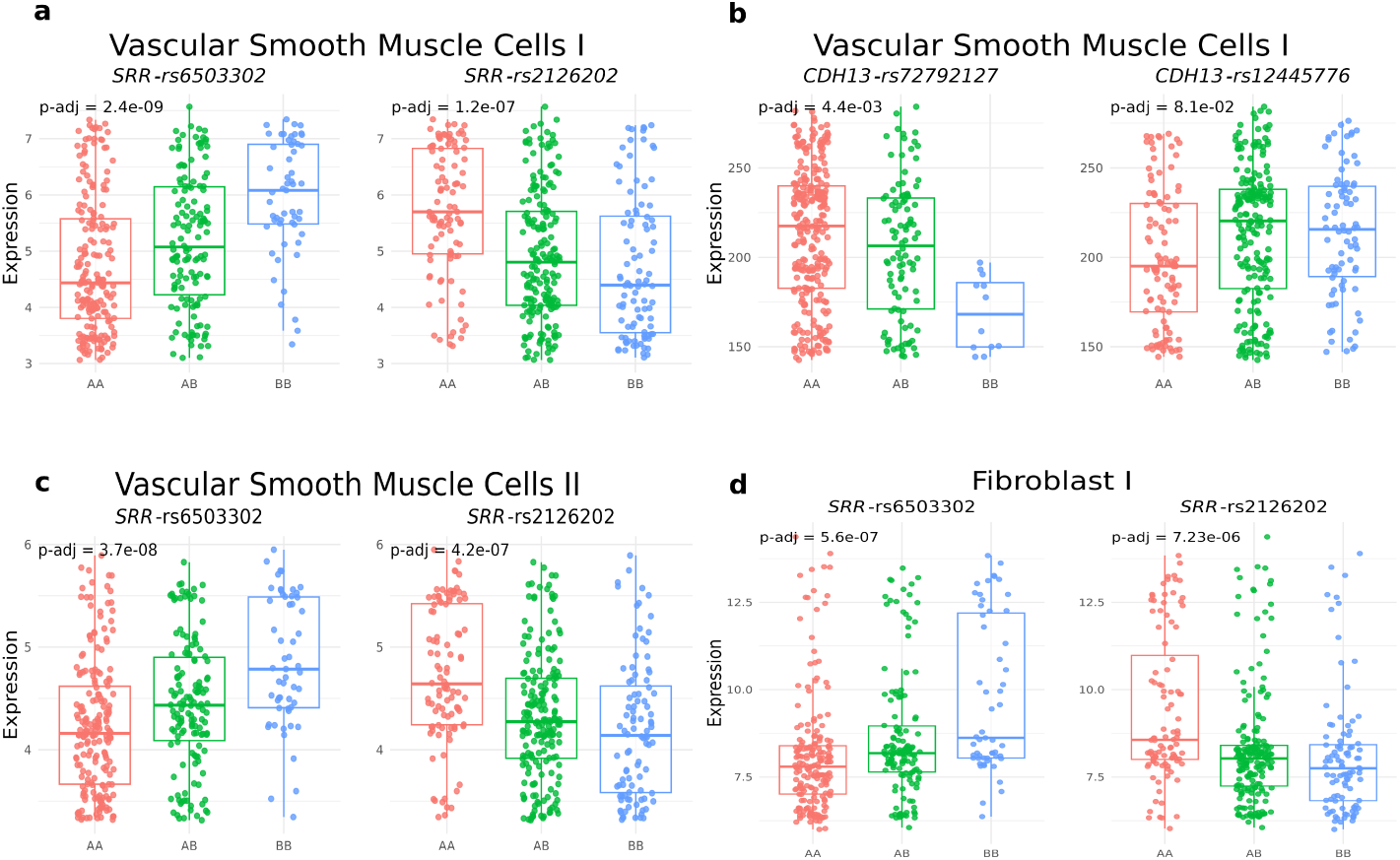
a-d) Boxplots with overlaid stripcharts for variants that showed a significant association (after correcting for sex and age) with the inferred expression of a-b) *SRR* and *CDH13* in VSMC I, c) *SRR* in VSMC II, and d) *SRR* in Fibroblast I. Genotypes are represented as AA, AB, and BB, on x-axis where AA denotes the absence of the effect allele, AB corresponds to heterozygotes, and BB indicates two copies of the effect allele (i.e. homozygosity for the effect allele).

## Discussion

The aortic wall comprises a diverse mix of cell types, each contributing uniquely to wall integrity, elasticity, and pulsatility. Aortic distensibility, or elasticity, decreases with age. The impact of age on distensibility would likely be more significant in a cohort with greater age variation than that found in the UK Biobank. Previous GWA studies have highlighted genetic variants associated with distensibility. Some single nucleotide polymorphisms (SNPs) associated with aortic distensibility have been reported to be expression quantitative trait loci (eQTLs); for instance, rs57130712 has been shown to significantly affect the expression of the *SRR* gene in arterial tissue [13]. Aortic stiffening, characterized by reduced distensibility and increased resistance to deformation, results from complex interactions among extracellular matrix components, including fibrillin, collagen, elastin fibers, and vascular smooth muscle cells[1]. Aortic diseases often involve the weakening and dilation of the aortic wall, typically accompanied by degradation in contractile smooth muscle cells (SMC) and the extracellular matrix (ECM) [18]. Despite these associations, the detailed molecular and cellular mechanisms driving changes in distensibility remain largely elusive. Thus, this study explored potential molecular mechanisms underlying both age-related and genetic variation in aortic distensibility.

Using gene expression deconvolution on GTEx aortic samples, we observed that the inferred proportion of vascular smooth muscle cells II increased and that pericytes and fibroblast I significantly decreased with age. Vascular smooth muscle cells, a key heterogenous cell type, provide structural and functional integrity of the aortic wall and synthesis of the extracellular matrix. Contractile SMCs (VSMC type I) help maintain vascular tone. On the contrary, an increase in synthetic smooth muscle cells (VSMC type II) deteriorates the aortic cell wall structure by reducing the expression of contraction-related genes [41]. Further, loss in pericytes is known to disrupt the vasa vasorum, which can lead to endothelial dysfunction, increased permeability, and impaired angiogenesis [42]. Reduction in pericytes can also contribute to degradation of cell repair processes [43]. Together with fibroblasts, pericytes can affect the ECM environment and influence the inflammatory environment [44].

By performing a cell-type-specific gene expression analysis, we found that the majority of gene expression correlations with age were in VSMC I, VSMC II, and Fibroblast I. The direction of correlation was the same for the majority of the genes across all cell types, indicating that age may impact gene expression in a comparable manner across cell types. Gene set enrichment analysis showed that genes in the GOBP gene sets *Increased Inflammatory Response* and *Abnormality Of Connective Tissue* were enriched among those whose expression decreased with age in VSMC I (Supplementary Figure 1). Overrepresentation analysis indicated that this downregulation may be influenced by negatively age-correlated genes. Additionally, the gene expression perturbation in the enriched *HP Abnormality Of Connective Tissue* gene set could impact extracellular matrix (ECM) production and degradation, potentially leading to vessel weakening. Furthermore, it was noteworthy to find enriched gene ontology biological process gene sets, such as *Vasculature Development* in VSMC II and *Regulation Of Membrane Potential* in VSMC I. These findings support our hypothesis that the change in genetic expression in specific cell types might affect the overall structural features of the aorta, which might lead to aortic wall abnormalities, such as loss of elasticity. However, further validation is needed to confirm the functional roles of these pathways.

Our study extends previous GWA studies of aortic distensibility (with 32,639 [39]; 29,895 [13]; 29,684 participants [40]) to a larger subset of the UK Biobank (47,674 participants). With this increased sample size, we identified four additional four loci (out of 12) associated with aortic distensibility; however, all of these loci had already been reported in earlier studies that incorporated multiple genetically correlated aortic phenotypes (Supplementary Table 1). Thus, while the larger dataset improved the power to detect associations, no completely novel loci were discovered, and our findings are consistent with the existing literature. As the aorta tends to lose its elasticity with age, we investigated whether genes mapped to SNPs associated with distensibility affect the extent to which aortic distensibility changes with age. Interestingly, the expression levels of 14 genes out of 27 mapped to GWAS loci demonstrated an association with age across at least one cell type. It was particularly intriguing that most of the distensibility associated genes showed significant associations in VSMC I and VSMC II (Figure 5-c). This observation prompted us to search for statistical interactions between distensibility-related SNPs and age. Two SNPs, rs10102879 and rs28545768, showed a significant interaction effect. We propose that these genetic variants may contribute to inter-individual variation in aortic distensibility by altering the rate at which the aorta stiffens with age.

We also assessed whether the genetic variants that were associated with distensibility in the GWAS were associated with the cell-type composition of aortic tissues in the GTEx samples. Although some genetic variations exhibited a nominally significant association with cell proportions, none survived FDR correction for multiple testing. However, we cannot rule out that some of the genetic associations observed may result from an effect of genetic variants on the relative proportion of different constituent cell types in the aorta as the modest number of samples in GTEx relative to the UKB dataset severely limits the power to detect such associations. To further investigate the impact of genetic variants, we conducted cell-type specific gene expression analysis. This aimed to identify variants associated with variation in gene expression within specific cell types and to evaluate whether variations in cell-type composition could contribute to the genetic variation in this important physiological phenotype. The GWAS loci were associated with the expression of genes *SRR* and *CDH13* in Vascular smooth muscle cells II, Fibroblast I, and Pericytes. Interestingly, *CDH13* was already known to play a role in angiogenesis that is involved in vasculature development, and previous studies suggest that across several cell types, it might be involved in aortic remodelling during aneurysmal degeneration [45].

Overall, this study integrates gene expression deconvolution and cell type-specific gene set analysis with genome wide association of aortic distensibility. Using gene expression deconvolution applied to aortic tissue samples, we identified significant age-related changes in the estimated proportions of Pericytes, VSMC II, and Fibroblast-I. We hypothesized that distensibility-associated SNPs might contribute to these cellular shifts. However, we did not detect significant associations between SNPs associated with aortic distensibility and cell type proportions following BH-based multiple test correction. We further explored whether these variants influence gene expression in specific aortic cell types. This cell-type-specific gene expression analysis highlighted two GWAS loci that had already been identified as eQTLs for *SRR* and *CDH13*. Our results add to these previous findings by providing evidence that these eQTLs specifically affect the expression of these two genes in VSMC I, VSMC II and Fibroblast I cells, shedding greater light on the potential mechanisms through which these loci exert an effect on aortic distensibility.

## Data Availability

UK Biobank data are not publicly available due to participant privacy and data-use restrictions. Access to the UK Biobank can be obtained by approved researchers through an application process (application number 74519). GTEx genotype and expression data were obtained through dbGaP under approved Project 20932. Access to GTEx controlled-access data requires an application to the dbGaP Data Access Committee.

## Acknowledgements

We thank the participants and coordinators of the UK Biobank for their invaluable contributions as this research was conducted using the UK Biobank Resource under Application Number 74519. We also acknowledge the participants of the Genotype-Tissue Expression (GTEx) Project, from whom genotypic as well as gene-expression data were obtained through dbGaP, with approval from the Data Access Committee under Project 20932.

We would also like to thank members of the Seoighe lab - Declan Bennett, Dónal O’Shea, Sophie Matthews, Harrison Anthony, Tyler Medina as well as Kevin Ryan, Barry Digby, Anthony Walters, Shir Dahan at the University of Galway; Ouso Daniel, Sodiq Ayobami Hameed at University College Dublin for valuable discussions regarding this research.

## Sources of funding

This publication has emanated from research conducted with the financial support of Research Ireland under grant number 18/CRT/6214. The funders had no role in study design, data collection and analysis, decision to publish, or preparation of the manuscript.

## Disclosures

The authors declare no competing interests.

## Notes

### Competing Interest Statement

The authors have declared no competing interest.

### Funding Statement

This publication has emanated from research conducted with the financial support of Research Ireland under grant number 18/CRT/6214. The funders had no role in study design, data collection and analysis, decision to publish, or preparation of the manuscript.

### Author Declarations

This research was conducted using the UK Biobank Resource under Application Number 74519. We also acknowledge the participants of the Genotype-Tissue Expression (GTEx) Project, from whom genotypic and gene-expression data were obtained through dbGaP, with approval from the Data Access Committee under Project 20932.

