## Supplementary for "Impact of cell type specific variations and age in aortic distensibility"

#### Polygenic Risk Score of Ascending distensibility

In a univariate regression, the polygenic score was a statistically significant predictor of the trait (p value = 3.75e-07), accounting for 0.1% of the variance ($R^2$ = 0.001). This effect size was markedly smaller than that of age and sex with $R^2$ of 0.27 (p value = 5.6e-07), which explained a substantially greater proportion of variance.

#### GWAS of descending distensibility

Similar to GWAS of ascending distensibility, an earlier GWAS was carried out on the majority of these samples (29,895; [1] 33,907; [2]). Using FUMA, these 29 independent loci could be mapped to 36 genes (Figure 1; Table 1). The overall SNP-heritability ($h^2g$) of descending aortic distensibility was estimated at 0.24, indicating that genetic factors account for a moderate portion of the variation observed.

#### Supplementary Figure legends

**Supplementary Figure 1:** a, b) Density plots illustrating the tendency for a more negative correlation with age for genes in the Human Phenotype Ontology gene sets Abnormality of Connective Tissue and Increased Inflammatory Response in VSMC I compared against the background genes using mremaR.

**Supplementary Figure 2:** Manhattan plot from the GWAS analysis of aortic descending distensibility. The x-axis represents chromosomes 1 to 22, and the y-axis represents the negative log (to the base 10) of the two-sided p values for the association of variants with ascending aortic distensibility.

**Supplementary Figure 3:** Intersection showing the overlap between the number of associated loci in Ascending (AAo) and Descending (DAo) distensibility. The table includes common loci and the nearest mapped gene.

**Supplementary Figure 4:** Venn diagram represents the intersection of the number of same associated independent SNPs in Ascending (in Red) and Descending distensibility (in Blue). The table contains common independent SNPs and the nearest mapped genes associated with ascending and descending distensibility.

**Supplementary Figure 5:** a-c) The top 3 associations of genetic variants with inferred cellular proportions (none of the variants survived FDR-based correction for multiple testing). Genotypes are represented as AA, AB, and BB, where AA denotes the absence of the effect allele, AB corresponds to heterozygotes, and BB indicates two copies of the effect allele (i.e. homozygosity for the effect allele).

#### Supplementary Table legends

**Supplementary Table 1:** Genomic locus blocks identified in the current genome-wide association analysis are shown together with regions previously reported using multi-trait analyses. For each locus, the lead SNP, its p value, and other independently significant variants detected in the current study are displayed. Loci marked with an asterisk (*) indicate regions whose lead SNP positions overlap with, or fall within, genomic intervals previously identified using BOLT-LMM. Earlier studies reported different lead SNPs within these regions, their correspondence to our loci is inferred based on chromosomal position and locus boundaries rather than exact SNP matches. Additional loci previously reported through multi trait analysis also fall within regions identified in our analysis of ascending aortic distensibility using BOLT-LMM.

**Supplementary Table 2:** Significant independent SNPs associated with descending distensibility using FUMA.

**Supplementary Table 3:** Independent SNPs (chromosome (chr) and base-pair position (BP) associated with ascending distensibility. The corresponding cell type (column 1) and significance levels are shown, with p values before and after Benjamini–Hochberg (BH) correction.

### Supplementary Tables

**Supplementary Table 1:** Genomic locus blocks identified in the current genome-wide association analysis are shown together with regions previously reported using multi-trait analyses. For each locus, the lead SNP, its p value, and other independently significant variants detected in the current study are displayed. Loci marked with an asterisk (*) indicate regions whose lead SNP positions overlap with, or fall within, genomic intervals previously identified using BOLT-LMM. Earlier studies reported different lead SNPs within these regions, their correspondence to our loci is inferred based on chromosomal position (chr:pos) and locus boundaries rather than exact SNP matches. Additional loci previously reported through multi trait analysis also fall within regions identified in our analysis of ascending aortic distensibility using BOLT-LMM.

| **Locus** | **Genomic Loci** | **Previously reported Genomic Locus [1]** | **Lead SNP rsID** | **p value** | **Individual Significant SNPs** |
|---|---|---|---|---|---|
| 1* | 3:41750260-42065005 | 3:41749669-42183563 | 3:41896441:T:TAA AAAAAAAAAAA | 1.70E-10 | 3:41896441_TAA AAAAAAAAAAA_T |
| 2 | 5:51135630-51200058 | 5:51158351-51201544 | rs4371706 | 2.50E-11 | rs4371706 |
| 3* | 5:95503950-95763256 | 5:95159822-95776105 | rs7447610 | 6.30E-13 | rs7447610; rs4336380 |
| 4 | 5:122489555-122556164 | 5:121855956-123164515 | rs337101 | 8.00E-12 | rs337101 |
| 5* | 7:73414589-73493533 | 7:7329388-73567718 | rs11768878 | 1.00E-31 | rs11768878; rs4717861; rs10224499; rs73144900; rs1859761; rs810549 |
| 6* | 8:75573675-75788406 | 8:75540855-75788406 | rs71527276 | 0.000000001 | rs71527276; rs7831476; rs141775964 |
| 6* | 8:75573675-75788406 | 8:75573675-75788406 | rs2732010 | 1.90E-11 | rs2732010; rs7831476; rs141775964 |
| 7* | 8:122628180-122702987 | 8:122627847-122734202 | rs10102879 | 1.00E-11 | rs10102879; rs11781907; 8:122674851_TTA_T; rs28545768 |
| 8* | 8:124541280-124610213 | 8:124541280-124615765 | rs10101794 | 0.000000011 | rs10101794; rs10086151; rs6470156 |
| 8* | 8:124541280-124610213 | 8:124541280-124610213 | rs34557926 | 1.50E-12 | rs34557926; rs10086151; rs6470156; rs11776999 |
| 9 | 10:30072785-30171017 | 10:30034986-30171504 | rs9336085 | 1.10E-10 | rs9336085 |
| 9 | 10:30072785-30171017 | 10:30034986-30171504 | rs914279 | 3.50E-12 | rs914279; rs2150562 |
| 10 | 10:95892706-95946164 | 10:95743087-97063487 | rs11187803 | 0.00000003 | rs11187803 |
| 11* | 16:82998900-83077384 | 16:83001098-83077384 | rs12445776 | 4.7E-09 | rs12445776; rs72792127 |
| 12* | 17:2015612-2220815 | 17:1931965-2333758 | rs216199 | 1.80E-11 | rs216199; rs6503302; rs2126202 |

**Supplementary Table 2:** Significant independent SNPs associated with descending distensibility using FUMA

| Locus | Independent SNP rsID | Chr | BP | p value | Mapped genes |
|---|---|---|---|---|---|
| 1 | rs11690961 | 2 | 46363336 | 3.30E-08 | *EPAS1, PRKCE* |
| 2 | rs73886131 | 3 | 186977783 | 9.10E-14 | *MASP1, SST, RTP2, RP11-211G3.3* |
| 2 | rs56210869 | 3 | 187004880 | 4.40E-08 | *MASP1, SST, RTP2, RP11-211G3.3* |
| 3 | rs6820391 | 4 | 54414696 | 1.90E-08 | *LNX1, CHIC2, FIP1L1, AC110792.1* |
| 4 | rs11766156 | 7 | 35528567 | 1.20E-08 | |
| 5 | rs3110697 | 7 | 45955029 | 7.80E-09 | *IGFBP3, AC011294.3* |
| 5 | 7:45964807_TGA_T | 7 | 45964807 | 1.80E-10 | *IGFBP3, AC011294.3* |
| 6 | 7:73426344_CG_C | 7 | 73426344 | 1.00E-12 | *ELN* |
| 7 | rs1449544 | 8 | 76591880 | 3.70E-10 | *ZFHX4* |
| 8 | rs2740776 | 8 | 92003130 | 1.80E-08 | *NECAB1, TMEM55A, OTUD6B, C8orf88* |
| 8 | rs28542498 | 8 | 92054476 | 2.10E-10 | *TMEM55A, C8orf88* |
| 8 | rs7832313 | 8 | 92198211 | 2.50E-09 | *NACAB1, SLC26A7, LRRC69, C8orf88* |
| 9 | rs10086151 | 8 | 124541280 | 2.00E-10 | *FBXO32* |
| 9 | rs10101794 | 8 | 124541764 | 9.20E-09 | *FBXO32* |
| 9 | rs6470155 | 8 | 124573833 | 1.00E-10 | *FBXO32* |

| Locus | Independent SNP rsID | Chr | BP | p value | Mapped genes |
|---|---|---|---|---|---|
| 9 | rs6470156 | 8 | 124579985 | 6.40E-15 | *FBXO32* |
| 9 | rs7814093 | 8 | 124595483 | 1.70E-08 | *FBXO32* |
| 9 | rs12547165 | 8 | 124605960 | 3.50E-12 | *FBXO32* |
| 9 | rs34557926 | 8 | 124607159 | 7.50E-17 | *FBXO32* |
| 10 | rs2202 | 10 | 30168699 | 1.20E-10 | *SVIL* |
| 11 | rs1343094 | 10 | 95900635 | 5.20E-17 | *PLCE1* |
| 11 | rs10882399 | 10 | 95903188 | 8.00E-15 | *PLCE1* |
| 11 | rs2274224 | 10 | 96039597 | 7.60E-10 | *TBC1D12, HELLS, PLCE1, NOC3L, SLC35G1* |
| 11 | rs2077218 | 10 | 96071561 | 5.20E-11 | *TBC1D12, HELLS, PLCE1, NOC3L, SLC35G1* |
| 11 | rs9420636 | 10 | 96132617 | 3.70E-09 | *TBC1D12, C10orf129, NOC3L, SLC35G1* |
| 11 | rs10430657 | 10 | 96451173 | 4.20E-09 | *TBC1D12, CYP2C18, HELLS, CYP2CP, CYP2C19, C10orf129, NOC3L, SLC35G1* |
| 12 | rs2244642 | 14 | 92359066 | 7.00E-11 | *CATSPERB, FBLN5* |
| 12 | rs8013684 | 14 | 92365790 | 2.40E-11 | *CATSPERB, FBLN5, TC2N* |
| 13 | rs72983206 | 19 | 11274065 | 5.60E-10 | *SPC24, KANK2* |

**Supplementary Table 3:** Independent SNPs (chromosome (chr) and base-pair position (BP)) associated with ascending distensibility. The corresponding cell type (column 1) and significance levels are shown, with p values before and after Benjamini–Hochberg (BH) correction.

| **Cell types** | **Chr** | **BP** | **p value** | **p-adj (BH)** |
|---|---|---|---|---|
| Fibroblast II | 8 | 74868420 | 0.00089 | 0.361 |
| VSMC I | 8 | 74782674 | 0.0091 | 0.437 |
| VSMC II | 8 | 123529524 | 0.0076 | 0.437 |
| Pericyte | 10 | 94184493 | 0.008 | 0.437 |
| Mesothelial | 8 | 74782674 | 0.009 | 0.437 |
| Fibroblast II | 8 | 74782674 | 0.00539 | 0.437 |
| Neuronal | 8 | 74844937 | 0.0093 | 0.437 |
| Neuronal | 8 | 74868420 | 0.0034 | 0.437 |
| Neuronal | 9 | 123529524 | 0.0068 | 0.437 |
| Fibroblast I | 8 | 123529524 | 0.012 | 0.481 |
| Endothelial II | 7 | 74040299 | 0.013144067 | 0.4815508 |
| Pericyte | 8 | 123529040 | 0.018208104 | 0.5241333 |
| Endothelial I | 5 | 96279156 | 0.018028072 | 0.5241333 |
| Lymphatic Endothelial | 8 | 74868420 | 0.016548273 | 0.5241333 |
| Fibroblast I | 8 | 74782674 | 0.019522333 | 0.5245 |
| Pericyte | 7 | 74009516 | 0.021121723 | 0.5320034 |
| Mesothelial | 8 | 74868420 | 0.024057896 | 0.5703137 |
| Mesothelial | 8 | 74844937 | 0.036629159 | 0.6161491 |
| Fibroblast II | 5 | 51889410 | 0.040606188 | 0.6161491 |
| Fibroblast I | 8 | 121669162 | 0.037790288 | 0.6161491 |
| Fibroblast I | 10 | 29794728 | 0.030119103 | 0.6161491 |
| Fibroblast I | 16 | 83003297 | 0.042004326 | 0.6161491 |
| Endothelial I | 5 | 96269410 | 0.045424425 | 0.6161491 |
| Endothelial I | 8 | 74782674 | 0.047396084 | 0.6161491 |
| Endothelial I | 8 | 123529524 | 0.039400973 | 0.6161491 |
| Lymphatic Endothelial | 8 | 74782674 | 0.035185783 | 0.6161491 |
| Lymphocyte | 8 | 74844937 | 0.046181774 | 0.6161491 |
| Lymphocyte | 8 | 74868420 | 0.043735025 | 0.6161491 |
| Neuronal | 7 | 74025935 | 0.0390009 | 0.6161491 |
| Neuronal | 8 | 123529040 | 0.042947705 | 0.6161491 |

### Supplementary Figures

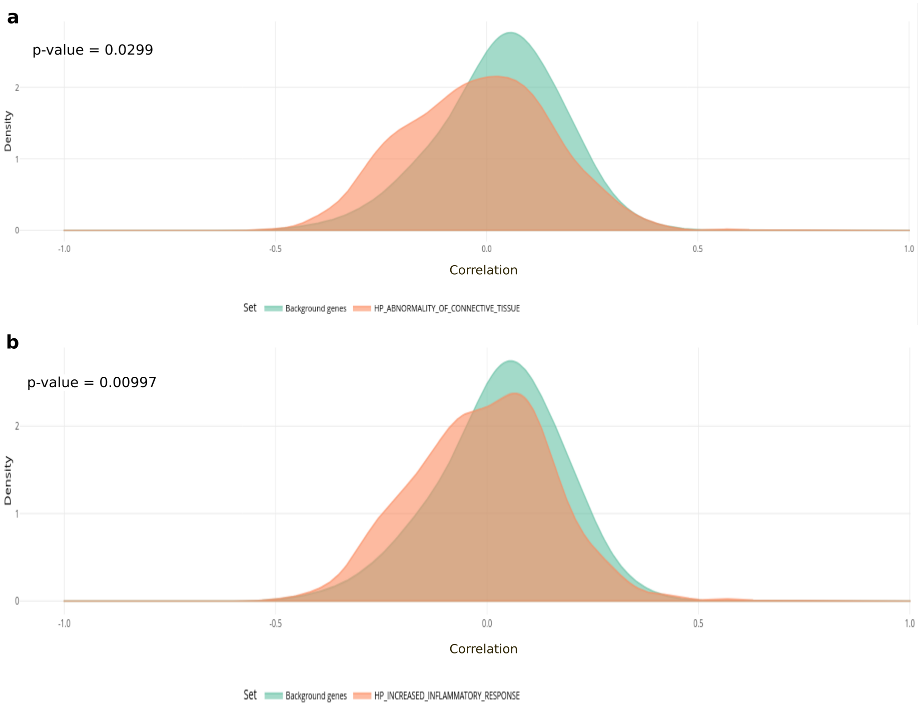


**Supplementary Figure 1: a, b)** Density plots illustrating the tendency for a more negative correlation with age for genes in the Human Phenotype Ontology gene sets Abnormality of Connective Tissue and Increased Inflammatory Response in VSMC I compared against the background genes using mremaR.

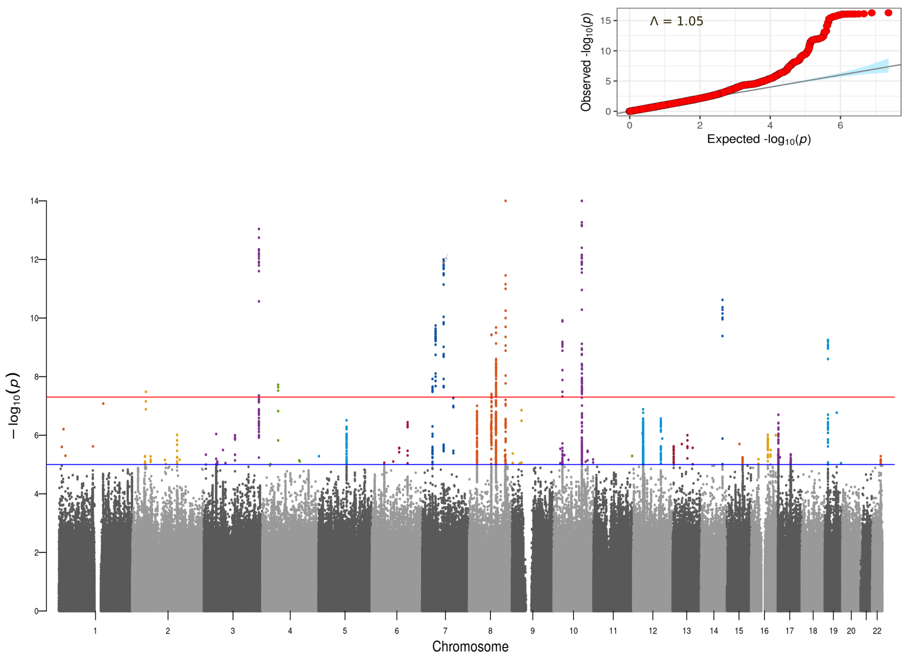


**Supplementary Figure 2:** Manhattan plot from the GWAS analysis of aortic descending distensibility. The x-axis represents chromosomes 1 to 22, and the y-axis represents the negative log (to the base 10) of the two-sided p values for the association of variants with ascending aortic distensibility.

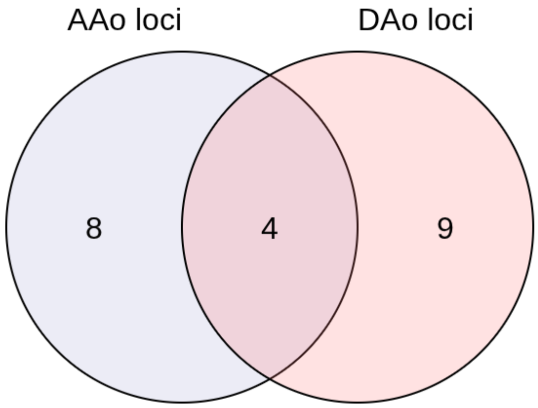


| AAo loci | DAo loci | Mapped gene |
|---|---|---|
| 7:73414589-73493533 | 7:73414589-73470714 | *ELN* |
| 8:124541280-124610213 | 8:124541280-124615765 | *FBXO32* |
| 10:30072785-30171017 | 10:30113768-30170487 | *SVIL* |
| 10:95892706-95946164 | 10:95892659-96712400 | *PLCE1* |

**Supplementary Figure 3:** The intersection shows the overlap between the number of associated loci in Ascending (AAo loci) and Descending (DAo loci) distensibility. The table includes common loci and the associated gene.

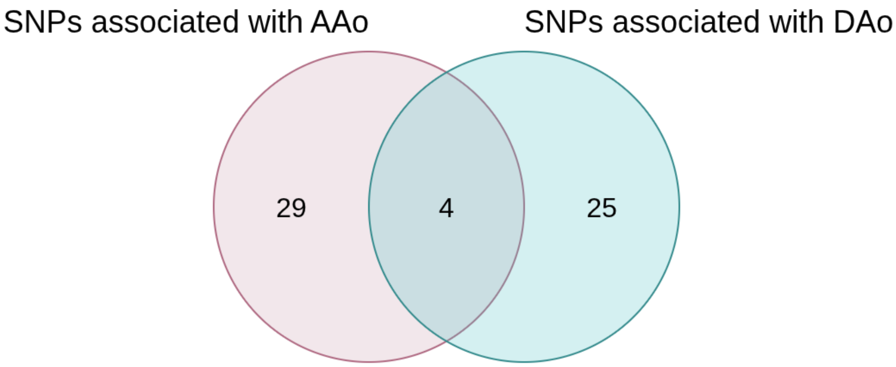


| rsid | Mapped gene |
|---|---|
| rs10086151 | *FBXO32* |
| rs10101794 | *FBXO32* |
| rs34557926 | *FBXO32* |
| rs6470156 | *FBXO32* |

**Supplementary Figure 4:** Venn diagram represents the intersection of the number of same associated independent SNPs in ascending (in Red) and descending distensibility (in Blue). The table contains common independent SNPs and the nearest mapped genes associated with ascending and descending distensibility.

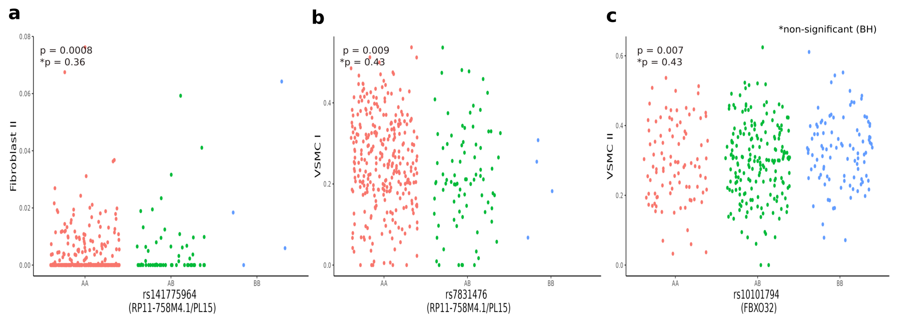


**Supplementary Figure 5:** a-c) The top 3 associations of genetic variants with inferred cellular proportions (none of the variants survived FDR-based correction for multiple testing). Genotypes are represented as AA, AB, and BB, where AA denotes the absence of the effect allele, AB corresponds to heterozygotes, and BB indicates two copies of the effect allele (i.e. homozygosity for the effect allele).
